# Analysis of Whole Genome Sequencing in a Cohort of Individuals with PHACE Syndrome Suggests Dysregulation of RAS/PI3K Signaling

**DOI:** 10.1101/2021.08.05.21261553

**Authors:** Elizabeth S. Partan, Francine Blei, Sarah L. Chamlin, Olivia M. T. Davies, Beth A. Drolet, Ilona J. Frieden, Ioannis Karakikes, Chien-Wei Lin, Anthony J. Mancini, Denise Metry, Anthony Oro, Nicole S. Stefanko, Laksshman Sundaram, Monika Tutaj, Alexander E. Urban, Kevin C. Wang, Xiaowei Zhu, Nara Sobreira, Dawn H. Siegel

## Abstract

The acronym PHACE stands for the co-occurrence of posterior brain fossa malformations, hemangiomas, arterial anomalies, cardiac defects, and eye abnormalities. The majority of patients have a segmental hemangioma and at least one developmental structural anomaly. The etiology and pathogenesis are unknown. Here we discuss the candidate causative genes identified in a *de novo* analysis of whole genome sequencing of germline samples from 98 unrelated trios in which the probands had PHACE, all sequenced as part of the Gabriella Miller Kids First Pediatric Research Program. A g:Profiler pathway analysis of the genes with rare, *de novo* variants suggested dysregulation of the RAS/MAPK and PI3K/AKT pathways that regulate cell growth, migration, and angiogenesis. These findings, along with the developmental anomalies and the vascular birthmark, support including PHACE within the RASopathy family of syndromes.

## INTRODUCTION

The RASopathies represent a collection of rare genetic conditions caused by mutations in individual genes within the RAS/MAP kinase (MAPK) pathway. RAS/MAPK signaling is upstream of many cellular pathways; therefore, depending on the individual mutation and affected tissue, single-gene perturbations can cause numerous syndromes, including Cardio-Facio-Cutaneous (CFC), Costello (CS), Legius (LS), Neurofibromatosis type 1 (NF1), Noonan (NS), Noonan-like (NS with Multiple Lentigines, NSML; NS with Loose Anagen Hair, NSLH) and Capillary Malformation-Arteriovenous Malformation (CM-AVM) (Grant et al. 2018; Stevenson et al. 2016). These syndromes share many clinical features including distinct facial features, developmental delays, cardiac defects, growth delays, and structural brain anomalies. While these individual syndromes are rare, as a group the RASopathies are among the most common genetic conditions in the world. The vast majority of mosaic neurocutaneous syndromes have been shown to be caused by somatic variants in RAS pathway genes, including the association of *HRAS* and *KRAS* variants with sebaceous nevus syndrome and *NRAS* variants with neurocutaneous melanosis (Chacon□macho et al. 2019; Kinsler et al. 2013).

The PHACE (OMIM 606519) acronym represents the sporadic vascular syndrome exhibiting the co-occurrence of posterior brain fossa malformations, hemangiomas, arterial anomalies, cardiac defects, and eye abnormalities (Frieden et al. 1996). The majority of patients have a segmental infantile hemangioma and at least one other feature (Garzon et al. 2016; Metry et al. 2009). Dysplasia and stenosis of the arteries of the head and neck is the second most common feature (occurring in >85%), followed by structural brain anomalies (including hypoplastic cerebellum), cardiovascular anomalies (including aortic coarctation or ventricular septal defect), and eye anomalies (Metry et al. 2009; Siegel 2017). The significant overlap in developmental anomaly phenotypes between PHACE and RASopathy patients suggests a common mechanistic underpinning of sporadic PHACE and germline RASopathy variants (Ruggieri et al. 2020).

The etiology and pathogenesis of PHACE are unknown, although consideration of the variety of body systems affected suggests that the age of onset is between 3 and 12 weeks of gestation during early vasculogenesis (Frieden et al. 1996; Metry et al. 2006; Siegel 2017). There is a 6:1 female:male preponderance, but no significant difference in the severity of the phenotype between males and females (Metry et al. 2009; Wan et al. 2017). PHACE is a sporadic disorder; there are no reported familial cases and at least four female patients with PHACE have given birth to clinically unaffected children (Martel et al. 2015; Stefanko et al. 2019). PHACE is believed to have a genetic cause because there is no evidence of a common exposure during gestation or early infancy for children diagnosed with PHACE (Wan et al. 2017). Autosomal dominant inheritance caused by *de novo*, germline pathogenic variants or mosaicism caused by somatic mosaic pathogenic variants have been suggested as possible modes of inheritance (Mitchell et al. 2012; Siegel 2017).

Here we present the *de novo* analysis of whole genome sequencing (WGS) of germline samples from 98 unrelated trios in which the probands had PHACE, all sequenced as part of the Gabriella Miller Kids First project (X01HL140519-01). Additionally, we analyzed the DECIPHER database (Firth et al. 2009) for genes affected by copy number variants (CNVs) identified in individuals with the search terms “hemangioma” and “facial hemangioma” and used this analysis to prioritize genes with *de novo* variants identified among our cohort of 98 patients. Among the rare, *de novo* variants, we prioritized variants in 10 genes in 18 patients; six of these genes were in the RAS pathway. Our results support including PHACE within the RASopathy family of syndromes.

## RESULTS

### Germline whole genome sequencing and *de novo* analysis pipeline

We identified a total of 16,107 *de novo* single nucleotide variants (SNVs) in our cohort of 98 patients; on average, this is 164 SNVs per patient. Our analysis found 15,752 rare (minor allele frequency [MAF] < 1%), *de novo* variants, including 219 coding variants (119 missense/nonsense/stoploss/splicing and 100 synonymous) and 15,533 noncoding variants. No *de novo* variant was found in more than two patients, and no gene had nonsynonymous, *de novo* coding variants in more than one patient.

### Variant prioritization

Among the rare, *de novo* variants, we prioritized 18 candidate SNVs in 10 genes in 18 patients, and two indels in these same genes (Supplemental Table 1; see Methods for a description of prioritization metrics). Briefly, variants were prioritized if they were rare or novel (population frequency < 1% or 0), affected a gene known to cause a phenotype similar to PHACE in humans or mice, or affected genes disrupted by CNVs in a previous study of PHACE (Siegel et al. 2013).

#### Genes associated with vascular human phenotypes

Using OMIM annotation, we identified a missense, heterozygous *STAMBP*–p.Ile301Asn variant in Patient 16. Missense and frameshift homozygous or compound heterozygous variants in this gene cause autosomal recessive microcephaly-capillary malformation syndrome (OMIM 614261), which is characterized by capillary malformation, microcephaly, optic atrophy, cleft palate, tapered fingers, and seizures. *STAMBP*–p.Ile301Asn is not found in gnomAD and is predicted to be pathogenic by 18 *in silico* algorithms. Additionally, 88% of non-VUS missense variants in this gene are pathogenic. Patient 16 was heterozygous for the *STAMBP*–p.Ile301Asn variant and a second candidate variant in this gene was not identified despite careful analysis of the WGS data including a search for deletions or duplications.

#### Genes associated with vascular mouse phenotypes

We then identified coding and noncoding variants in six genes associated with lethal knockout mouse models due to vascular abnormalities. Two variants, *RASA3*–p.Val85Met in Patient 4 and *THBS2*–p.Asp859Asn in Patient 14, were exonic. The *RASA3*– p.Val85Met variant is predicted to be pathogenic by 11 algorithms (CADD, DEOGEN2, EIGEN, FATHMM-MKL, FATHMM-XF, LRT, Mutation assessor, MutationTaster, PrimateAI, SIFT, and SIFT4G) and to affect splicing. The *THBS2*–p.Asp859Asn variant is found in the C-terminal type 3 repeats and likely disrupts intraprotein interactions and calcium binding (Figure 1); this variant is predicted to be pathogenic by 19 algorithms (BayesDel-addAF, BayesDel-noAF, CADD, DEOGEN2, EIGEN, EIGEN PC, FATHMM, FATHMM-MKL, FATHMM-XF, LIST-S2, MVP, MetaLR, MetaSVM, Mutation assessor, MutationTaster, PROVEAN, REVEL, SIFT, and SIFT4G). The other variants were noncoding and included two intronic SNVs in *BCAS3*, three intronic SNVs in *DLC1*, one intronic SNV in *GLRX3*, one intronic SNV in *PIK3CA*, and one intronic SNV in *THBS2* (see Supplemental Table 1). Most of the identified variants are novel. Variants in *BCAS3*, *DLC1*, *GLRX3*, and *PIK3CA* are predicted to affect splicing. Variants in *BCAS3*, *DLC1*, *PIK3CA*, and *THBS2* are predicted to be pathogenic by at least one algorithm. The *BCAS3* SNV in intron 23 is predicted to fall within a binding site for GATA1 and the *THBS2* intronic SNV is predicted to fall in a binding site for IKZF1. All three variants in *DLC1* and the variants in *GLRX3* and *PIK3CA* are predicted to be in open chromatin in human vascular endothelial cells (HUVECs) by Encode and therefore accessible to proteins regulating gene expression: four are predicted to be transcribed, and one (*DLC1* intron 5) is predicted to be in an enhancer.

**Figure 1:**
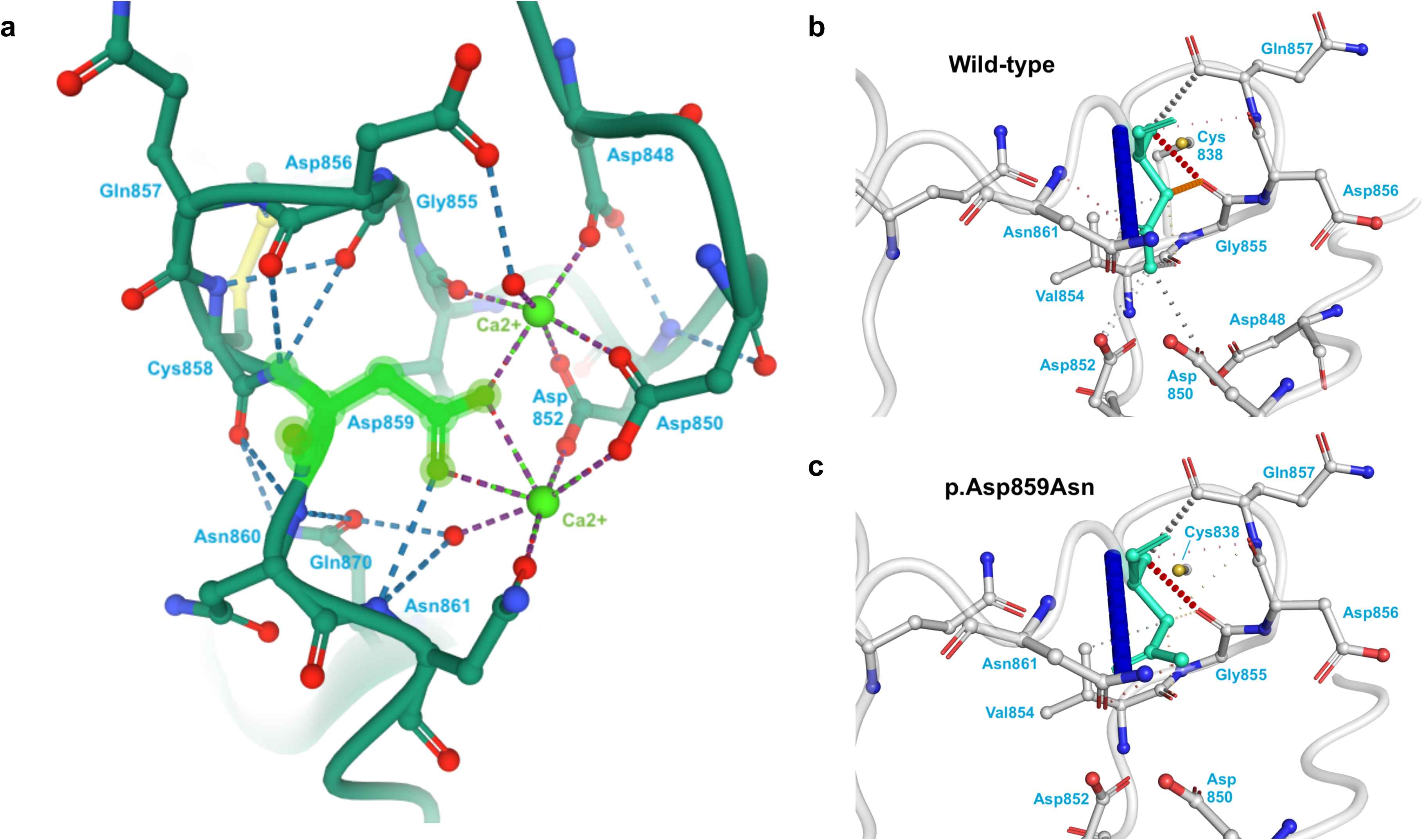
Location and consequences of *THBS2*–p.Asp859Asn variant. **A)** The location and interactions of p.Asp859 in the THBS2 protein structure 1YO8 in PDB. The p.Asp859 amino acid is highlighted in green. The surrounding amino acids that participate in hydrogen binding with p.Asp859 and/or coordination with the calcium ions are labeled. **B-C)** The wild-type **(B)** and mutant **(C)** structure predictions from DynaMut are given. The affected amino acid is highlighted in teal. Red dashes indicate hydrogen bonds, and orange dashes indicate weak hydrogen bonds. The p.Asp859Asn variant disrupts many of the interactions of p.Asp859 with its neighboring amino acids.

#### Genes previously implicated in CNV analysis

We next looked for variants in genes affected by CNVs identified in a cohort of patients with PHACE described by Siegel et al. (2013) among our cohort. This analysis identified the aforementioned intronic variant in *PIK3CA* again. We also prioritized two intronic variants in *AFF2*, an intronic variant in *EPHA3*, and four intronic variants in *EXOC4*. All four genes were affected by a CNV in a single patient in the prior study. The *EPHA3* intronic variant is predicted to be pathogenic by EIGEN (0.58), to affect splicing, and to fall within binding sites for CTCF, TCF12, and RAD21. All four *EXOC4* variants are predicted to affect splicing, and the *EXOC4* variant in intron 17 is predicted to be pathogenic by EIGEN (0.05) and GWAVA (0.5) and to fall within binding sites for GATA2 and GATA3.

#### Indels in prioritized genes

Finally, we investigated our cohort for rare, *de novo* indels affecting any of the 12 prioritized genes and identified two more variants: a 2-bp intronic deletion in *BCAS3* in Patient 20 and an intronic 8-bp insertion in *THBS2* in Patient 12, just upstream of the previously identified intronic SNV in *THBS2* in the same patient.

### DECIPHER analyses

Next, we analyzed published CNVs using DECIPHER to identify previously reported individuals with a phenotypic description containing the term “hemangioma” and CNVs containing our prioritized genes. This analysis identified five individuals in DECIPHER with deletions affecting *THBS2*; a single individual each with a deletion affecting *BCAS3*, *GLRX3*, and *RASA3*; and a single individual each with a duplication affecting *DLC1* and *PIK3CA*.

We expanded our DECIPHER analysis to identify individuals with “facial hemangioma” and the genes disrupted by their CNVs. Next, we searched for rare, *de novo* variants in our cohort in the genes disrupted in two or more DECIPHER individuals. *THBS2* was only disrupted by deletions in DECIPHER. Another two genes were each disrupted by two CNVs, one deletion and one duplication, in DECIPHER individuals with facial hemangioma. *ALDH3A1* has a novel p.Val373Met variant in our Patient 8; this SNV is in the first codon of exon eight, is predicted to affect splicing by Human Splicing Finder (HSF), and is predicted to be pathogenic by CADD (23.3) and FATHMM-MKL (0.48). *ATP7B* has a p.Thr482Ala variant in our Patient 10; this SNV is predicted to affect splicing by HSF.

We performed an enrichment analysis for the DECIPHER CNVs disrupting *ALDH3A1*, *ATP7B*, and *THBS2*. Only deletions disrupting *THBS2* were significantly more common among individuals with facial hemangioma (FET, p=3.7e-2, OR=7.17) or hemangioma (FET, p=3.9e-3, OR=5.24) in DECIPHER than among the individuals without these features in DECIPHER, respectively.

### Pathway analysis

Genes containing rare, *de novo*, non-intergenic SNVs (n=4,320 genes) were analyzed through g:Profiler to look for common pathways disrupted in our cohort (Table 1). The five gene ontology (GO) terms with the most significant p-values were all related to development and morphogenesis. The most significant term, nervous system development (adjusted p=6.22e-24), included our candidate genes *AFF2*, *DLC1*, *EPHA3*, *PIK3CA*, and *THBS2*. We also observed an enrichment for cell adhesion (adjusted p=1.34e-10, including *BCAS3*, *DLC1*, *EPHA3*, *PIK3CA*, and *THBS2*), which is critical for angiogenesis. The three KEGG terms with the most significant p-values implicated RAP signaling (adjusted p=1.61e-6, including *PIK3CA*), focal adhesion (adjusted p=2.43e-6, including *PIK3CA* and *THBS2*), and phospholipase D signaling (adjusted p=8.13e-6, including *PIK3CA*).

**Table 1:** g:Profiler pathway analysis of genes with rare, de novo variants.

A) Gene ontology (GO) terms
| Term ID | Term name | Adjusted p-value | # genes |
| --- | --- | --- | --- |
| GO:0007399 | nervous system development | 6.22E-24 | 646 |
| GO:0000902 | cell morphogenesis | 1.88E-23 | 329 |
| GO:0032989 | cellular component morphogenesis | 6.85E-20 | 260 |
| GO:0048666 | neuron development | 8.51E-20 | 343 |
| GO:0032990 | cell part morphogenesis | 1.15E-19 | 236 |
| GO:0016043 | cellular component organization | 1.84E-19 | 1413 |
| GO:0031175 | neuron projection development | 4.82E-19 | 309 |
| GO:0030182 | neuron differentiation | 5.53E-19 | 401 |
| GO:0120039 | plasma membrane bounded cell projection morphogenesis | 6.95E-19 | 228 |
| GO:0120036 | plasma membrane bounded cell projection organization | 1.54E-18 | 428 |
| GO:0048858 | cell projection morphogenesis | 1.60E-18 | 228 |
| GO:0048812 | neuron projection morphogenesis | 2.63E-18 | 223 |
| GO:0048699 | generation of neurons | 1.10E-17 | 433 |
| GO:0030030 | cell projection organization | 2.90E-17 | 432 |
| GO:0071840 | cellular component organization or biogenesis | 7.38E-17 | 1432 |
| GO:0043167 | ion binding | 1.10E-16 | 1378 |
| GO:0048468 | cell development | 1.23E-16 | 563 |
| GO:0000904 | cell morphogenesis involved in differentiation | 1.31E-16 | 239 |
| GO:0022008 | neurogenesis | 4.92E-16 | 449 |
| GO:0048667 | cell morphogenesis involved in neuron differentiation | 3.46E-15 | 199 |
| GO:0051128 | regulation of cellular component organization | 6.13E-15 | 599 |
| GO:0019899 | enzyme binding | 9.62E-15 | 566 |
| GO:0010975 | regulation of neuron projection development | 1.34E-14 | 176 |
| GO:0034330 | cell junction organization | 4.11E-14 | 217 |
| GO:0048856 | anatomical structure development | 5.63E-14 | 1312 |
| GO:0051960 | regulation of nervous system development | 6.34E-14 | 280 |
| GO:0120035 | regulation of plasma membrane bounded cell projection organization | 8.37E-14 | 217 |
| GO:0031344 | regulation of cell projection organization | 2.28E-13 | 218 |
| GO:0007275 | multicellular organism development | 4.63E-13 | 1211 |
| GO:0045664 | regulation of neuron differentiation | 6.17E-13 | 211 |
| GO:0022604 | regulation of cell morphogenesis | 6.51E-13 | 166 |
| GO:0009653 | anatomical structure morphogenesis | 7.53E-13 | 669 |
| GO:0048731 | system development | 5.70E-12 | 1101 |
| GO:0032502 | developmental process | 5.86E-12 | 1397 |
| GO:0043168 | anion binding | 4.70E-11 | 666 |
| GO:0007264 | small GTPase mediated signal transduction | 9.61E-11 | 162 |
| GO:0050767 | regulation of neurogenesis | 1.28E-10 | 243 |
| GO:0007155 | cell adhesion | 1.34E-10 | 375 |
| GO:0034329 | cell junction assembly | 1.51E-10 | 142 |
| GO:0022610 | biological adhesion | 1.70E-10 | 376 |
| GO:0060284 | regulation of cell development | 2.17E-10 | 273 |
| GO:0051130 | positive regulation of cellular component organization | 3.48E-10 | 320 |
| GO:0016358 | dendrite development | 4.05E-10 | 94 |
| GO:0023051 | regulation of signaling | 4.09E-10 | 820 |
| GO:0046872 | metal ion binding | 1.06E-09 | 930 |
| GO:0051056 | regulation of small GTPase mediated signal transduction | 1.09E-09 | 112 |
| GO:0007010 | cytoskeleton organization | 1.28E-09 | 356 |
| GO:0050839 | cell adhesion molecule binding | 1.78E-09 | 163 |
| GO:0065008 | regulation of biological quality | 2.06E-09 | 899 |
| GO:0030029 | actin filament-based process | 2.57E-09 | 226 |
| GO:0010769 | regulation of cell morphogenesis involved in differentiation | 3.24E-09 | 108 |
| GO:0010646 | regulation of cell communication | 3.33E-09 | 806 |
| GO:0043169 | cation binding | 3.43E-09 | 943 |
| GO:0050773 | regulation of dendrite development | 3.93E-09 | 66 |
| GO:0050808 | synapse organization | 6.33E-09 | 133 |
| GO:0007409 | axonogenesis | 6.46E-09 | 150 |
| GO:0006996 | organelle organization | 9.75E-09 | 877 |
| GO:0032559 | adenyl ribonucleotide binding | 1.99E-08 | 383 |
| GO:0031346 | positive regulation of cell projection organization | 2.79E-08 | 127 |
| GO:0030554 | adenyl nucleotide binding | 4.80E-08 | 383 |
| GO:0051020 | GTPase binding | 5.13E-08 | 162 |
| GO:0019904 | protein domain specific binding | 5.21E-08 | 198 |
| GO:0044093 | positive regulation of molecular function | 5.34E-08 | 436 |
| GO:0005524 | ATP binding | 6.16E-08 | 368 |
| GO:0030036 | actin cytoskeleton organization | 6.68E-08 | 198 |
| GO:0061564 | axon development | 8.00E-08 | 157 |
| GO:0010976 | positive regulation of neuron projection development | 8.83E-08 | 100 |
| GO:0051962 | positive regulation of nervous system development | 9.77E-08 | 165 |
| GO:0099536 | synaptic signaling | 1.00E-07 | 200 |
| GO:0007169 | transmembrane receptor protein tyrosine kinase signaling pathway | 2.31E-07 | 207 |
| GO:0017016 | Ras GTPase binding | 2.65E-07 | 127 |
| GO:0099537 | trans-synaptic signaling | 3.45E-07 | 193 |
| GO:0097367 | carbohydrate derivative binding | 4.49E-07 | 521 |
| GO:0051179 | localization | 4.95E-07 | 1402 |
| GO:0031267 | small GTPase binding | 5.62E-07 | 129 |
| GO:0005085 | guanyl-nucleotide exchange factor activity | 7.05E-07 | 78 |
| GO:0008092 | cytoskeletal protein binding | 7.54E-07 | 252 |
| GO:0035556 | intracellular signal transduction | 8.74E-07 | 632 |
| GO:0022607 | cellular component assembly | 9.90E-07 | 658 |
| GO:0032879 | regulation of localization | 1.04E-06 | 643 |
| GO:0030165 | PDZ domain binding | 1.11E-06 | 40 |
| GO:0045666 | positive regulation of neuron differentiation | 1.32E-06 | 120 |
| GO:0043087 | regulation of GTPase activity | 1.85E-06 | 141 |
| GO:0048522 | positive regulation of cellular process | 2.05E-06 | 1190 |
| GO:0044087 | regulation of cellular component biogenesis | 2.11E-06 | 252 |
| GO:0097485 | neuron projection guidance | 2.18E-06 | 94 |
| GO:0098916 | anterograde trans-synaptic signaling | 2.20E-06 | 188 |
| GO:0007268 | chemical synaptic transmission | 2.20E-06 | 188 |
| GO:0007416 | synapse assembly | 2.75E-06 | 67 |
| GO:0140096 | catalytic activity, acting on a protein | 2.88E-06 | 512 |
| GO:0032553 | ribonucleotide binding | 3.02E-06 | 446 |
| GO:0005088 | Ras guanyl-nucleotide exchange factor activity | 3.18E-06 | 48 |
| GO:0032555 | purine ribonucleotide binding | 3.67E-06 | 442 |
| GO:0007411 | axon guidance | 4.05E-06 | 93 |
| GO:0043085 | positive regulation of catalytic activity | 4.06E-06 | 352 |
| GO:0050804 | modulation of chemical synaptic transmission | 4.09E-06 | 125 |
| GO:0005509 | calcium ion binding | 4.56E-06 | 191 |
| GO:0099177 | regulation of trans-synaptic signaling | 4.80E-06 | 125 |
| GO:0017076 | purine nucleotide binding | 5.98E-06 | 443 |
| GO:0005515 | protein binding | 6.42E-06 | 2726 |
| GO:0004672 | protein kinase activity | 7.71E-06 | 163 |
| GO:0005543 | phospholipid binding | 8.46E-06 | 130 |
| GO:0016773 | phosphotransferase activity, alcohol group as acceptor | 8.79E-06 | 186 |
| GO:0048813 | dendrite morphogenesis | 1.23E-05 | 57 |
| GO:0008047 | enzyme activator activity | 1.41E-05 | 146 |
| GO:0009966 | regulation of signal transduction | 1.45E-05 | 697 |
| GO:0006928 | movement of cell or subcellular component | 1.72E-05 | 514 |
| GO:0035639 | purine ribonucleoside triphosphate binding | 1.78E-05 | 424 |
| GO:0007417 | central nervous system development | 2.20E-05 | 263 |
| GO:0003779 | actin binding | 2.23E-05 | 125 |
| GO:0004674 | protein serine/threonine kinase activity | 2.32E-05 | 127 |
| GO:1900006 | positive regulation of dendrite development | 2.51E-05 | 35 |
| GO:0000166 | nucleotide binding | 2.83E-05 | 486 |
| GO:1901265 | nucleoside phosphate binding | 3.01E-05 | 486 |
| GO:0007167 | enzyme linked receptor protein signaling pathway | 3.04E-05 | 270 |
| GO:0050769 | positive regulation of neurogenesis | 4.05E-05 | 140 |
| GO:0050770 | regulation of axonogenesis | 4.21E-05 | 66 |
| GO:0003824 | catalytic activity | 4.34E-05 | 1198 |
| GO:1901888 | regulation of cell junction assembly | 4.39E-05 | 68 |
| GO:0006793 | phosphorus metabolic process | 4.54E-05 | 731 |
| GO:0050807 | regulation of synapse organization | 5.70E-05 | 72 |
| GO:0060589 | nucleoside-triphosphatase regulator activity | 6.63E-05 | 102 |
| GO:0007420 | brain development | 6.98E-05 | 201 |
| GO:0006796 | phosphate-containing compound metabolic process | 7.81E-05 | 724 |
| GO:0043547 | positive regulation of GTPase activity | 8.09E-05 | 118 |
| GO:0007015 | actin filament organization | 8.49E-05 | 125 |
| GO:0031589 | cell-substrate adhesion | 8.93E-05 | 107 |
| GO:0016301 | kinase activity | 9.70E-05 | 204 |
| GO:0050803 | regulation of synapse structure or activity | 1.07E-04 | 74 |
| GO:0017124 | SH3 domain binding | 1.17E-04 | 48 |
| GO:0099173 | postsynapse organization | 1.31E-04 | 56 |
| GO:0036094 | small molecule binding | 1.59E-04 | 562 |
| GO:0099003 | vesicle-mediated transport in synapse | 2.05E-04 | 64 |
| GO:0007265 | Ras protein signal transduction | 2.14E-04 | 102 |
| GO:0065009 | regulation of molecular function | 2.14E-04 | 658 |
| GO:0033036 | macromolecule localization | 2.22E-04 | 689 |
| GO:0016772 | transferase activity, transferring phosphorus-containing groups | 2.31E-04 | 235 |
| GO:0032535 | regulation of cellular component size | 2.34E-04 | 112 |
| GO:0060322 | head development | 2.38E-04 | 208 |
| GO:0030695 | GTPase regulator activity | 2.70E-04 | 91 |
| GO:0007267 | cell-cell signaling | 3.04E-04 | 407 |
| GO:0046578 | regulation of Ras protein signal transduction | 3.20E-04 | 65 |
| GO:0010720 | positive regulation of cell development | 3.53E-04 | 155 |
| GO:0005096 | GTPase activator activity | 3.71E-04 | 83 |
| GO:0007215 | glutamate receptor signaling pathway | 3.83E-04 | 41 |
| GO:0032501 | multicellular organismal process | 4.13E-04 | 1568 |
| GO:0048638 | regulation of developmental growth | 4.16E-04 | 104 |
| GO:0044085 | cellular component biogenesis | 4.75E-04 | 683 |
| GO:0008289 | lipid binding | 4.80E-04 | 194 |
| GO:0005089 | Rho guanyl-nucleotide exchange factor activity | 4.82E-04 | 27 |
| GO:0098609 | cell-cell adhesion | 4.87E-04 | 218 |
| GO:0099504 | synaptic vesicle cycle | 6.06E-04 | 60 |
| GO:0031325 | positive regulation of cellular metabolic process | 6.53E-04 | 728 |
| GO:0048518 | positive regulation of biological process | 6.85E-04 | 1278 |
| GO:0007610 | behavior | 6.94E-04 | 160 |
| GO:0010770 | positive regulation of cell morphogenesis involved in differentiation | 7.11E-04 | 55 |
| GO:0035091 | phosphatidylinositol binding | 7.32E-04 | 78 |
| GO:0050793 | regulation of developmental process | 7.36E-04 | 605 |
| GO:0090066 | regulation of anatomical structure size | 7.48E-04 | 142 |
| GO:0019900 | kinase binding | 7.78E-04 | 188 |
| GO:0008104 | protein localization | 8.26E-04 | 609 |
| GO:0018193 | peptidyl-amino acid modification | 8.42E-04 | 305 |
| GO:0044089 | positive regulation of cellular component biogenesis | 8.53E-04 | 147 |
| GO:0051239 | regulation of multicellular organismal process | 8.55E-04 | 721 |
| GO:0061572 | actin filament bundle organization | 9.13E-04 | 56 |
| GO:0033043 | regulation of organelle organization | 9.80E-04 | 306 |
| GO:0050790 | regulation of catalytic activity | 1.10E-03 | 520 |
| GO:0019901 | protein kinase binding | 1.14E-03 | 169 |
| GO:0051173 | positive regulation of nitrogen compound metabolic process | 1.16E-03 | 690 |
| GO:0016740 | transferase activity | 1.27E-03 | 513 |
| GO:0003013 | circulatory system process | 1.33E-03 | 166 |
| GO:0040011 | locomotion | 1.60E-03 | 444 |
| GO:0036211 | protein modification process | 1.61E-03 | 876 |
| GO:0006464 | cellular protein modification process | 1.61E-03 | 876 |
| GO:0097435 | supramolecular fiber organization | 1.78E-03 | 182 |
| GO:2000026 | regulation of multicellular organismal development | 1.78E-03 | 491 |
| GO:0048583 | regulation of response to stimulus | 1.79E-03 | 899 |
| GO:0005178 | integrin binding | 1.82E-03 | 50 |
| GO:0051963 | regulation of synapse assembly | 1.92E-03 | 39 |
| GO:0051049 | regulation of transport | 2.04E-03 | 417 |
| GO:0030154 | cell differentiation | 2.20E-03 | 896 |
| GO:0048869 | cellular developmental process | 2.20E-03 | 911 |
| GO:0048589 | developmental growth | 2.26E-03 | 173 |
| GO:0048588 | developmental cell growth | 2.26E-03 | 74 |
| GO:0033674 | positive regulation of kinase activity | 2.57E-03 | 158 |
| GO:0051234 | establishment of localization | 2.59E-03 | 1089 |
| GO:0006468 | protein phosphorylation | 2.63E-03 | 447 |
| GO:0051017 | actin filament bundle assembly | 2.76E-03 | 54 |
| GO:0051345 | positive regulation of hydrolase activity | 2.94E-03 | 191 |
| GO:0042391 | regulation of membrane potential | 2.96E-03 | 121 |
| GO:0022603 | regulation of anatomical structure morphogenesis | 3.22E-03 | 282 |
| GO:0008361 | regulation of cell size | 3.28E-03 | 60 |
| GO:0016310 | phosphorylation | 3.46E-03 | 535 |
| GO:0005488 | binding | 3.68E-03 | 3124 |
| GO:0009893 | positive regulation of metabolic process | 3.71E-03 | 815 |
| GO:1990138 | neuron projection extension | 4.17E-03 | 57 |
| GO:0010604 | positive regulation of macromolecule metabolic process | 4.40E-03 | 757 |
| GO:0098742 | cell-cell adhesion via plasma-membrane adhesion molecules | 4.54E-03 | 81 |
| GO:0046873 | metal ion transmembrane transporter activity | 4.71E-03 | 116 |
| GO:0006935 | chemotaxis | 5.27E-03 | 165 |
| GO:0051641 | cellular localization | 5.39E-03 | 722 |
| GO:0031646 | positive regulation of nervous system process | 5.44E-03 | 28 |
| GO:0032970 | regulation of actin filament-based process | 5.97E-03 | 112 |
| GO:0042330 | taxis | 6.39E-03 | 165 |
| GO:0034220 | ion transmembrane transport | 6.77E-03 | 272 |
| GO:0015020 | glucuronosyltransferase activity | 6.93E-03 | 17 |
| GO:0019902 | phosphatase binding | 7.35E-03 | 60 |
| GO:0044260 | cellular macromolecule metabolic process | 7.38E-03 | 1639 |
| GO:0060627 | regulation of vesicle-mediated transport | 7.97E-03 | 140 |
| GO:0048814 | regulation of dendrite morphogenesis | 8.38E-03 | 36 |
| GO:0048041 | focal adhesion assembly | 8.46E-03 | 33 |
| GO:0048639 | positive regulation of developmental growth | 8.67E-03 | 60 |
| GO:0051094 | positive regulation of developmental process | 8.79E-03 | 334 |
| GO:0051493 | regulation of cytoskeleton organization | 9.17E-03 | 144 |
| GO:0048013 | ephrin receptor signaling pathway | 9.39E-03 | 34 |
| GO:0044877 | protein-containing complex binding | 9.50E-03 | 287 |
| GO:0042578 | phosphoric ester hydrolase activity | 9.92E-03 | 102 |
| GO:0048523 | negative regulation of cellular process | 1.00E-02 | 1019 |
| GO:1902414 | protein localization to cell junction | 1.02E-02 | 39 |
| GO:0010810 | regulation of cell-substrate adhesion | 1.03E-02 | 66 |
| GO:0034332 | adherens junction organization | 1.12E-02 | 28 |
| GO:0017048 | Rho GTPase binding | 1.12E-02 | 51 |
| GO:0060341 | regulation of cellular localization | 1.22E-02 | 235 |
| GO:0004115 | 3',5'-cyclic-AMP phosphodiesterase activity | 1.29E-02 | 9 |
| GO:0023056 | positive regulation of signaling | 1.33E-02 | 417 |
| GO:0006810 | transport | 1.63E-02 | 1056 |
| GO:1990782 | protein tyrosine kinase binding | 1.68E-02 | 35 |
| GO:0007156 | homophilic cell adhesion via plasma membrane adhesion molecules | 1.77E-02 | 54 |
| GO:0051347 | positive regulation of transferase activity | 1.84E-02 | 171 |
| GO:0010771 | negative regulation of cell morphogenesis involved in differentiation | 1.92E-02 | 36 |
| GO:0052696 | flavonoid glucuronidation | 1.95E-02 | 8 |
| GO:0034765 | regulation of ion transmembrane transport | 2.01E-02 | 128 |
| GO:0007160 | cell-matrix adhesion | 2.08E-02 | 69 |
| GO:0043412 | macromolecule modification | 2.14E-02 | 904 |
| GO:0010647 | positive regulation of cell communication | 2.19E-02 | 414 |
| GO:0060560 | developmental growth involved in morphogenesis | 2.21E-02 | 71 |
| GO:0040007 | growth | 2.34E-02 | 233 |
| GO:0008015 | blood circulation | 2.35E-02 | 142 |
| GO:0099175 | regulation of postsynapse organization | 2.36E-02 | 35 |
| GO:0006836 | neurotransmitter transport | 2.49E-02 | 65 |
| GO:0008081 | phosphoric diester hydrolase activity | 2.54E-02 | 33 |
| GO:0032956 | regulation of actin cytoskeleton organization | 2.88E-02 | 99 |
| GO:0040008 | regulation of growth | 3.00E-02 | 169 |
| GO:0007044 | cell-substrate junction assembly | 3.23E-02 | 36 |
| GO:0051258 | protein polymerization | 3.42E-02 | 84 |
| GO:0061387 | regulation of extent of cell growth | 3.55E-02 | 39 |
| GO:0004114 | 3',5'-cyclic-nucleotide phosphodiesterase activity | 3.61E-02 | 14 |
| GO:0008066 | glutamate receptor activity | 3.61E-02 | 14 |
| GO:0071495 | cellular response to endogenous stimulus | 3.93E-02 | 317 |
| GO:1902903 | regulation of supramolecular fiber organization | 4.07E-02 | 101 |
| GO:0045296 | cadherin binding | 4.16E-02 | 88 |
| GO:0016049 | cell growth | 4.29E-02 | 126 |
| GO:0045665 | negative regulation of neuron differentiation | 4.29E-02 | 67 |
| GO:0150115 | cell-substrate junction organization | 4.30E-02 | 37 |
| GO:0008038 | neuron recognition | 4.33E-02 | 22 |
| GO:0051336 | regulation of hydrolase activity | 4.41E-02 | 292 |
| GO:0017137 | Rab GTPase binding | 4.43E-02 | 52 |
| GO:0030010 | establishment of cell polarity | 4.50E-02 | 46 |
| GO:0005201 | extracellular matrix structural constituent | 4.60E-02 | 51 |
| GO:0051640 | organelle localization | 4.69E-02 | 160 |
| GO:0045595 | regulation of cell differentiation | 4.95E-02 | 423 |

B) KEGG pathways
| Term ID | Term name | Adjusted p-value | # genes |
| --- | --- | --- | --- |
| KEGG:04015 | Rap1 signaling pathway | 1.61E-06 | 72 |
| KEGG:04510 | focal adhesion | 2.43E-06 | 69 |
| KEGG:04072 | phospholipase D signaling pathway | 8.13E-06 | 54 |
| KEGG:04360 | axon guidance | 1.38E-05 | 62 |
| KEGG:04724 | glutamatergic synapse | 2.67E-05 | 44 |
| KEGG:04935 | growth hormone synthesis, secretion and action | 8.32E-05 | 44 |
| KEGG:05032 | morphine addiction | 8.98E-05 | 36 |
| KEGG:04211 | longevity regulating pathway | 2.05E-04 | 35 |
| KEGG:04213 | longevity regulating pathway - multiple species | 2.81E-04 | 27 |
| KEGG:04810 | regulation of actin cytoskeleton | 3.47E-04 | 66 |
| KEGG:04976 | bile secretion | 3.78E-04 | 35 |
| KEGG:04012 | ErbB signaling pathway | 5.57E-04 | 33 |
| KEGG:04512 | ECM-receptor interaction | 6.19E-04 | 34 |
| KEGG:04151 | PI3K-Akt signaling pathway | 9.34E-04 | 97 |
| KEGG:04022 | cGMP-PKG signaling pathway | 1.23E-03 | 53 |
| KEGG:04728 | dopaminergic synapse | 2.00E-03 | 44 |
| KEGG:01522 | endocrine resistance | 2.03E-03 | 35 |
| KEGG:04611 | platelet activation | 2.46E-03 | 42 |
| KEGG:04725 | cholinergic synapse | 2.98E-03 | 39 |
| KEGG:04928 | parathyroid hormone synthesis, secretion and action | 3.61E-03 | 37 |
| KEGG:04540 | gap junction | 4.93E-03 | 32 |
| KEGG:04730 | long-term depression | 5.70E-03 | 24 |
| KEGG:04713 | circadian entrainment | 6.84E-03 | 34 |
| KEGG:04020 | calcium signaling pathway | 6.91E-03 | 59 |
| KEGG:04911 | insulin secretion | 7.87E-03 | 31 |
| KEGG:04926 | relaxin signaling pathway | 1.31E-02 | 41 |
| KEGG:05100 | bacterial invasion of epithelial cells | 1.40E-02 | 27 |
| KEGG:05412 | arrhythmogenic right ventricular cardiomyopathy | 1.48E-02 | 28 |
| KEGG:04915 | estrogen signaling pathway | 1.52E-02 | 43 |
| KEGG:04720 | long-term potentiation | 2.12E-02 | 25 |
| KEGG:04520 | adherens junction | 2.25E-02 | 26 |
| KEGG:04723 | retrograde endocannabinoid signaling | 2.43E-02 | 45 |
| KEGG:04971 | gastric acid secretion | 3.04E-02 | 27 |
| KEGG:04062 | chemokine signaling pathway | 4.03E-02 | 54 |
| KEGG:04070 | phosphatidylinositol signaling system | 4.05E-02 | 32 |
| KEGG:04912 | GnRH signaling pathway | 4.07E-02 | 31 |
| KEGG:04925 | aldosterone synthesis and secretion | 4.99E-02 | 32 |

### Expression analysis

We explored the expression of the candidate genes *RASA3*, *AFF2*, *DLC1*, *EPHA3*, *PIK3CA*, and *THBS2* across diverse cell types in the human developing heart (Figure 2) by incorporating chromatin accessibility (scATAC-seq) (Ameen & Sundaram et. al., *unpublished*) and gene expression (scRNA-seq) datasets (Asp et al. 2019; Miao et al. 2020; Suryawanshi et al. 2020). We observed that *AFF2*, *EPHA3*, *PIK3CA*, and *THBS2* are co-expressed in the vasculature (pre-smooth muscle cells and vascular smooth muscle cells) in the fetal heart, whilst *AFF2*, *EPHA3*, and *PIK3CA* are co-expressed in the endothelium. Furthermore, *RASA3*, *DLC1*, and *THBS2* are co-expressed in pericytes, the mural cells surrounding blood vessels and embedded within the basement membrane of the vasculature and adjacent to endothelial cells (Armulik et al. 2005), and *RASA3* and *THBS2* are co-expressed in perivascular fibroblasts, which may serve as pericyte precursors (Rajan et al. 2020). Together, these data suggest that the expression of the candidate genes is enriched in endothelial cells and mural cells (pericytes and vascular smooth muscle cells) in the blood vessel wall.

**Figure 2:**
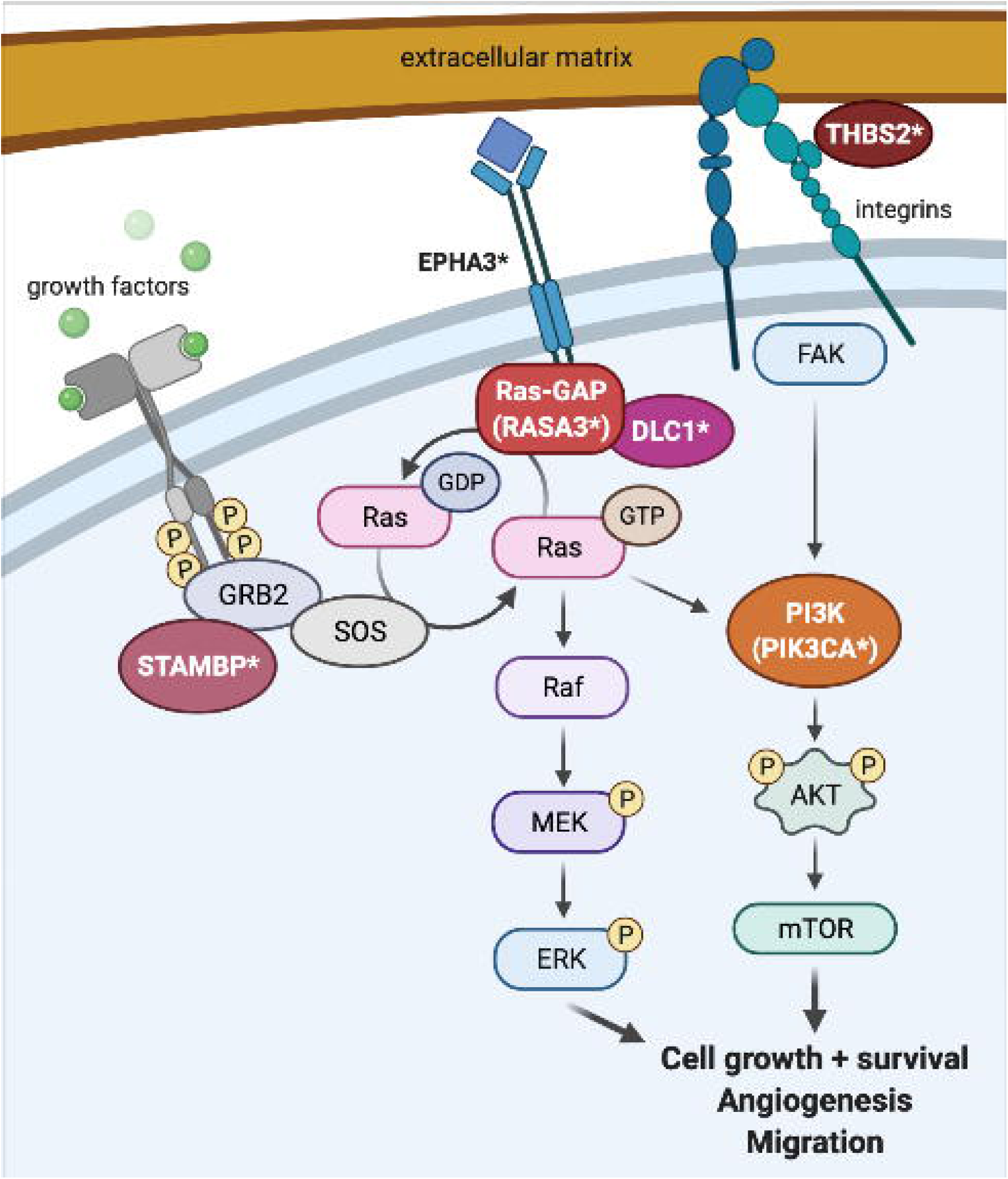
Expression of candidate genes in the developing heart at single cell level. **A-F)** Clustering of single cells by Uniform Manifold Approximation and Projection (UMAP) based on the differentially accessibility of open chromatin regions (scATAC-seq). Gene expression levels of *RASA3* **(A)**, *DLC1* **(B)**, *AFF2* **(C)**, *EPHA3* **(D)**, *PIK3CA* **(E)**, and *THBS2* **(F)** imputed from gene accessibility scores (scATAC-seq) and RNA expression (scRNA-seq). Annotation shows the expression level of each gene in the corresponding cell type cluster.

## DISCUSSION

Our whole genome sequencing analyses support a disruption in RAS pathway signaling as the primary pathogenic mechanism behind PHACE. The RAS pathway genes identified in our analysis are associated with abnormal angiogenesis, including changes to cytoskeletal organization, cell migration, and adhesion, in relevant knockout mouse models. DLC1 binds RASA1 (a paralog of RASA3) at focal adhesions, and *Dlc1* knockout causes embryonic lethal placenta vascular malformations (Durkin et al. 2005; Yang et al. 2009). EPHA3 regulates cytoskeletal organization, cell-cell adhesion, and cell migration in angiogenesis, and *Epha3* knockout causes perinatal lethal cardiac defects (Clifford et al. 2008; Vaidya et al. 2003; Vearing et al. 2005). PIK3CA regulates cell migration and adhesion and has been associated with vascular anomalies, and *Pik3ca* knockout causes embryonic lethal vascular defects and hemorrhage (Bi et al. 1999; Graupera et al. 2008; Lelievre et al. 2005). RASA3 regulates cell-cell and cell-matrix adhesion and cell migration, and *Rasa3* knockout causes embryonic lethal hemorrhage following vascular lumenization defects (Iwashita et al. 2007; Molina-Ortiz et al. 2018). THBS2 binds heparin in the extracellular matrix (ECM), mediates cell-matrix interactions, and inhibits cell adhesion and angiogenesis, and *Thbs2* knockout causes increased vascularity and premature death (Bornstein et al. 2000; Kyriakides et al. 1998; Noh et al. 2003; Streit et al. 1999; Yang et al. 2000).

The g:Profiler pathway analysis of the genes with rare, *de novo* SNVs in these patients also implicated these same candidate genes. KEGG pathway analysis identified RAP signaling, focal adhesion, and phospholipase D (PLD) signaling as the most significantly affected pathways. RASA3 downregulates RAP1 GTPase signaling (Battram et al. 2017; Molina-Ortiz et al. 2018). BCAS3, DLC1, PIK3CA, and THBS2 all participate in the regulation of focal adhesion (Durkin et al. 2005; Elzie and Murphy-Ullrich 2004; Jain et al. 2012; Matsuoka et al. 2012; Yang et al. 2009). PLD is required for actin cytoskeleton remodeling in cell migration and angiogenesis, participates in RAS activation, and is activated by both RAS/MAPK/ERK and PI3K/AKT/mTOR signaling (Bruntz et al. 2014). RASA3 downregulates RAS signaling through MEK/ERK and also binds PI3K and regulates cell migration though PI3K-dependent integrin signaling (Battram et al. 2017; Castellano and Downward 2011). PIK3CA forms part of the PI3K complex and regulates endothelial cell migration during vascular development (Graupera et al. 2008). STAMBP binds GRB2 to regulate both the RAS/MAPK and PI3K/AKT/mTOR pathways (McDonell et al. 2013; Tsang et al. 2006). THBS2 signals through FAK activation of the MAPK/ERK and PI3K/AKT pathways to stimulate actin reorganization, decrease cell adhesion, and increase migration (Goicoechea et al. 2000; Greenwood et al. 1998; Orr et al. 2002). DLC1 and EPHA3 both bind RASA1, a close paralog of RASA3 (Haupaix et al. 2013; Yang et al. 2009); the *EPHA3* intronic variant in particular is also in a binding site for TCF12, which downregulates ERK signaling (Yi et al. 2017). DLC1 is also phosphorylated by AKT, which abrogates its tumor suppression activity (Ko et al. 2010). The identification of variants in these genes among the probands with PHACE suggests that defects in matricellular signaling affecting endothelial cell adhesion and migration through the RAS and PI3K pathways may contribute to the pathogenesis of PHACE (Figure 3).

**Figure 3:**
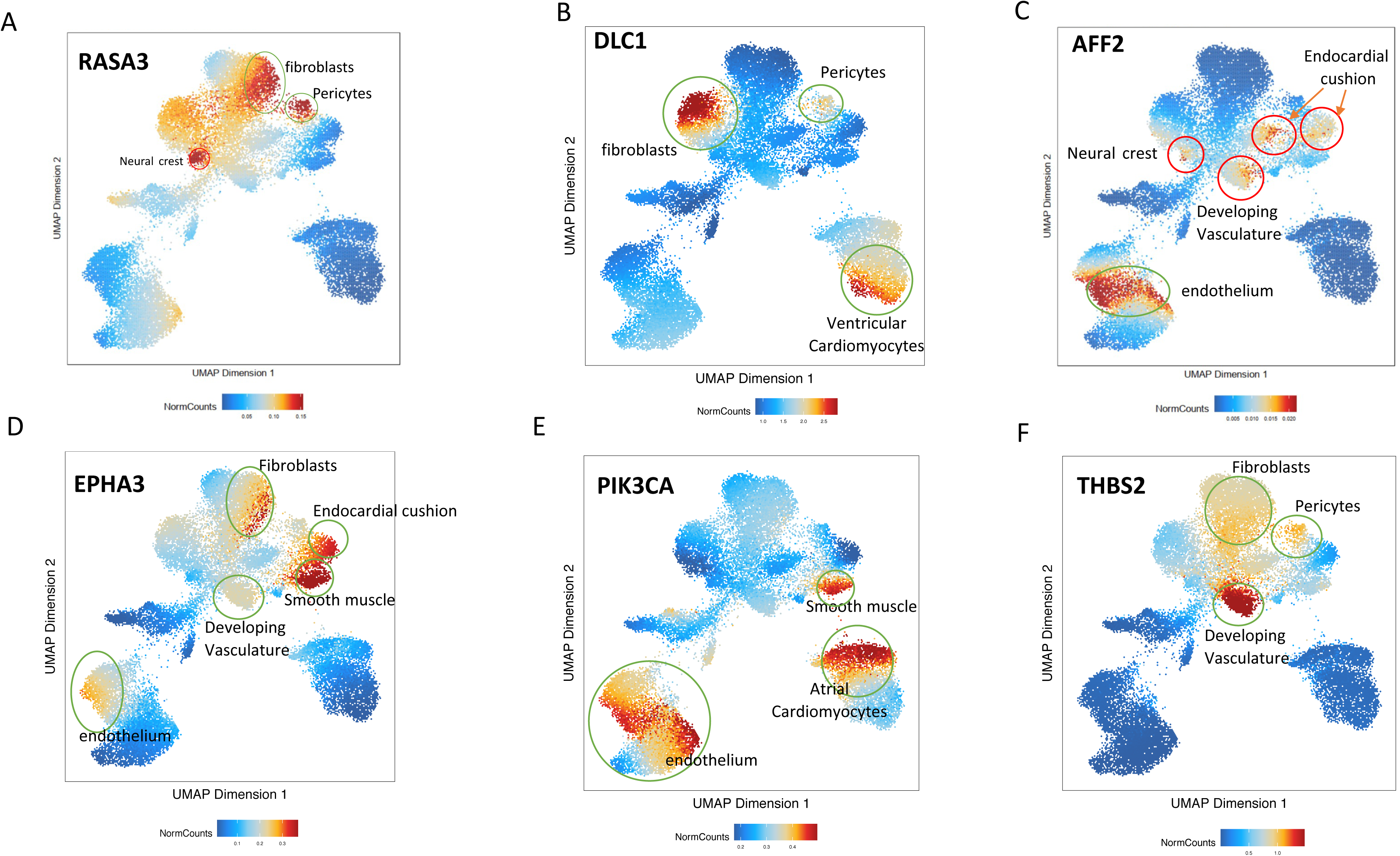
Diagram of proposed RAS signaling pathway. RAS is activated downstream of growth factor binding to cellular receptors, which signal through GRB2/SOS/STAMBP, and deactivated by RAS-GAP proteins (RASA1, RASA3), which interact with EPHA3 and DLC1. Activated RAS-GTP signals through the MEK/ERK and PI3K/AKT/mTOR pathways to upregulate cell growth, cell migration, and angiogenesis. PI3K is also activated through extracellular matrix (ECM) signaling: THBS2 binds to the ECM and to integrins, which link the cell to the ECM, and THBS2 signals through focal adhesion kinase (FAK) to PI3K. Protein names denoted with an asterisk (DLC1, EPHA3, PIK3CA, RASA3, STAMBP, THBS2) were found to have *de novo* variants in our cohort. Image created using BioRender.

Our analysis of rare, *de novo* germline variants combined with the DECIPHER analysis of CNVs found in individuals with hemangioma identified *THBS2* as a strong candidate causative gene for PHACE. *THBS2* had missense and intronic variants in our cohort and was deleted in five individuals with hemangioma in DECIPHER; an enrichment analysis showed that deletions disrupting *THBS2* were significantly more common among individuals with hemangioma than in individuals without this feature in DECIPHER. While no human phenotype has been associated with this gene, knockout of this gene in a mouse causes blood vessel overgrowth and premature death (Bornstein et al. 2000; Kyriakides et al. 1998; Yang et al. 2000). *THBS2* is highly expressed in arterial tissues (aorta, coronary, and tibial) described in GTEx and in the developing vasculature in our scRNA analysis. THBS2 interacts with the ECM, mediates cell-matrix interactions, inhibits cell adhesion, inhibits angiogenesis, and acts as a tumor suppressor (Bornstein et al. 2000; Kyriakides et al. 1998; Noh et al. 2003; Streit et al. 1999; Yang et al. 2000). The *THBS2* residue p.Asp859, which had a missense variant in our cohort, is found in the C-terminal type 3 repeats and participates in calcium binding (Figure 1). This calcium binding is necessary for correct folding of the C terminus, which itself must correctly trimerize in order to support the integrin-binding and cell motility functions of the THBS2 protein (Anilkumar et al. 2002; Carlson et al. 2008; Kvansakul et al. 2004). Therefore, a variant in this position could affect the protein function by disrupting protein folding and trimerization.

Our analysis identified *RASA3* as a second strong candidate causative gene for PHACE. *RASA3* had a missense variant in our cohort and was deleted in one DECIPHER individual with capillary hemangioma. Germline pathogenic variants in a related gene, *RASA1*, are known to cause capillary malformation–arteriovenous malformation (Eerola et al. 2003). No human phenotype has been associated with *RASA3*, but knockout of this gene in a mouse causes hemorrhage and embryonic lethality due to vascular abnormalities (Iwashita et al. 2007; Molina-Ortiz et al. 2018). *RASA3* also regulates angiogenesis via the RAS/RAP pathway (Molina-Ortiz et al. 2018), which was implicated in our pathway analysis, and was highly expressed in pericytes in our scRNA analysis.

We also identified seven noncoding variants in four RAS pathway genes (*DLC1*, *EPHA3*, *PIK3CA*, and *THBS2*). These variants were predicted to affect splicing, functional DNA, transcription factor binding, and/or open chromatin in vascular cells (Table 2). The genes *DLC1* and *PIK3CA* are highly intolerant to loss-of-function coding mutations (probability of loss-of-function intolerance scores > 0.9) (Lek et al. 2016) and have enhanced expression in PHACE-relevant tissues: *DLC1* in neurons and retinal amacrine cells and *PIK3CA* in aorta and coronary artery (Genotype-Tissue Expression (GTEx) Project 2021). Single cell RNA expression analyses also showed that our candidate RAS pathway genes have high expression in neural crest, smooth muscle, and endothelial cells in the developing heart (Figure 2). The predicted disruption of expression and/or splicing of these genes by our identified noncoding variants may therefore contribute to other PHACE features. For instance, an intronic SNV, rs1905014-T, in *DLC1* is significantly associated with optic disc size (Han et al. 2019). However, the effects of the noncoding variants on splicing and/or functional DNA need to be investigated further.

**Table 2:** Candidate germline *de novo* variants in RAS pathway genes.

| Gene | Patient ID | Patient phenotype | Variant (hg38) | Variant consequence | gnomAD MAF | Predicted effect on splicing? | Damaging functional DNA scores? | TFBS? | Open chromatin in HUVECs? | OMIM phenotype | Homozygous knockout mouse model | CNV(s) |
| --- | --- | --- | --- | --- | --- | --- | --- | --- | --- | --- | --- | --- |
| *DLC1* | Patient 1 | Infantile hemangioma; Cerebellar hypoplasia; Migration anomaly of cortex; Middle cerebral artery hypoplasia; Chorio-retinal coloboma | 8:13089422: C:T | intronic SNV (intron 15) | 1.9e-3 | — | — | — | transcriptional elongation | colorectal cancer, somatic | neural tube, heart, brain, and vascular placenta defects; embryonic lethality<sup>1</sup> | 1 duplication in DECIPHER [ID: 395935] |
|  | Patient 17 | Infantile hemangioma; Coarctation of aorta; Sternal pit | 8:13104433: G:A | intronic SNV (intron 7) | novel | — | EIGEN=0.25 | — | transcriptional elongation |  |  |  |
|  | Patient 3 | Infantile hemangioma; Internal carotid artery hypoplasia; Internal carotid artery tortuosity; Absent vertebral artery; Tortuous aorta with pseudo-coarctation | 8:13253055: G:C | intronic SNV (intron 5) | novel | HSF: activates cryptic acceptor site | — | — | strong enhancer |  |  |  |
| *EPHA3* | Patient 9 | Infantile hemangioma; Sternal cleft; Dental enamel defects | 3:89246570: G:A | intronic SNV (intron 3) | novel | HSF: activates cryptic acceptor site, changes exonic splicing silencer / enhancer | EIGEN=0.58 | CTCF, TCF12, RAD21 | — | — | perinatal death due to cardiac failure<sup>2</sup> | 1 duplication in PHACE |
| *PIK3CA* | Patient 5 | Infantile hemangioma; Subglottic hemangioma; Supra-umbilical raphe | 3:179200365: T:G | intronic SNV (intron 3) | novel | HSF: changes exonic splicing silencer / enhancer | EIGEN=0.21 | — | weakly transcribed | CLAPO syndrome, somatic; CLOVE syndrome, somatic; Cowden syndrome; keratosis, seborrheic, somatic; macrodactyly, somatic; megalencephaly-capillary malformation-polymicrogyria syndrome, somatic; nevus, epidermal, somatic; variety of cancers, somatic | vascular defects; hemorrhage; growth retardation; embryonic lethality<sup>3</sup> | 1 duplication in PHACE; 1 duplication in DECIPHER [ID: 283584] |

<sup>1</sup> (Durkin et al. 2005)
<sup>2</sup> (Vaidya et al. 2003)
<sup>3</sup> (Bi et al. 1999; Graupera et al. 2008; Lelievre et al. 2005)

| Gene | Patient ID | Patient phenotype | Variant (hg38) | Variant consequence | gnomAD MAF | Predicted effect on splicing? | Damaging functional DNA scores? | TFBS? | Open chromatin in HUVECs? | OMIM phenotype | Homozygous knockout mouse model | CNV(s) |
| --- | --- | --- | --- | --- | --- | --- | --- | --- | --- | --- | --- | --- |
| *RASA3* | Patient 4 | Hypoplasia of cerebellum; Infantile hemangioma; Internal carotid artery hypoplasia; Internal carotid artery tortuosity; Optic nerve dysplasia | 13:114041023: C:T | p.Val85Met | 1.2e-5 | HSF: changes exonic splicing silencer / enhancer | CADD=23.6; FATHMM=0.95; GWAVA=0.44 | — | transcriptional elongation | — | hemorrhage; embryonic lethality<sup>4</sup> | 1 deletion in DECIPHER [ID: 393047] |
| *STAMBP* | Patient 16 | Infantile hemangioma; Abnormal cerebral arterial morphology; Ventricular septal defect (VSD); Dental enamel hypoplasia | 2:73850410: T:A | p.Ile301Asn | novel | — | CADD=27.7; FATHMM=0.99 | — | weakly transcribed | microcephaly-capillary malformation syndrome | postnatal growth retardation; limb-clasping; hypocellular cerebral cortex; loss of hippocampal neurons; blepharoptosis; death of starvation at weaning<sup>5</sup> | — |
| *THBS2* | Patient 14 | Infantile hemangioma; Absent left internal carotid artery | 6:169225343: C:T | p.Asp589Asn | novel | — | CADD=28.4; FATHMM=0.96 | — | — | — | blood vessel abnormalities; premature death<sup>6</sup> | 5 deletions in DECIPHER [IDs: 392300<sup>7</sup>, 392970<sup>8</sup>, 392995<sup>9</sup>, 396392<sup>10</sup>, 402744] |
|  | Patient 12 | Infantile hemangioma; Aberrant right subclavian artery; Fetal origin of cerebral artery; Internal carotid artery hypoplasia; Internal carotid artery tortuosity; Vertebral artery hypoplasia; Apical muscular VSD | 6:169243142: C: CCACATTCT | intronic 8-bp insertion (intron 5) | novel | — | — | IKZF1 | — |  |  |  |
|  |  |  | 6:169243176: C:T | intronic SNV (intron 5) | 3.3e-5 | — | FATHMM=0.41 | IKZF1 | — |  |  |  |

<sup>4</sup> (Iwashita et al. 2007)
<sup>5</sup> (Ishii et al. 2001)
<sup>6</sup> (Kyriakides et al. 1998)
<sup>7</sup> (Peeden et al. 1983)
<sup>8</sup> (Römke et al. 1987)
<sup>9</sup> (Petit et al. 1980)
<sup>10</sup> (Hopkin et al. 1997)

In conclusion, we have found strong candidate coding and noncoding variants in genes in the RAS/MAPK and PI3K/AKT pathways in individual patients with PHACE. These variants, paired with the vascular birthmark (infantile hemangioma) and the structural brain, heart, arterial, and eye anomalies in PHACE, support the inclusion of PHACE as a RASopathy syndrome. It is striking that the majority of these variants were predicted to have effects that may lead to altered gene expression during development. However, because of the complex cellular interactions between vascular cell types, further *in vivo* studies are necessary to confirm pathogenicity of the variants identified and to understand their effect on the pathways that are likely disrupted by these variants.

## SUBJECTS AND METHODS

### Subjects

Patients with PHACE and their parents were recruited and sequenced as part of the Gabriella Miller Kids First Pediatric Research Program (X01HL140519-01). The study is approved by the Medical College of Wisconsin IRB #PRO00034867 and patients gave written, informed consent. Of the 98 patients discussed here, 19 were male, and 85 were white (Table 3). The average age of the cohort was 5.8 years at the time of enrollment. Ninety patients had facial hemangioma.

**Table 3:** Patient demographics.

|  | Number of patients |
| --- | --- |
| Gender |  |
| • Female | 79 |
| • Male | 19 |
| Race |  |
| • White | 85 |
| • Black or African American | 4 |
| • Asian | 2 |
| • American Indian or Alaska Native | 2 |
| • Other or Unknown | 5 |
| Ethnicity |  |
| • Not Hispanic or Latino | 88 |
| • Hispanic or Latino | 10 |
| Facial hemangioma | 90 |
| Age (average) | 5.8 years |

### Germline whole genome sequencing and *de novo* analysis pipeline

WGS was performed on germline DNA from 98 unrelated patients with PHACE and their parents. WGS libraries were sequenced on the Illumina NovaSeq 6000 platform using 150-bp paired ends and the NovaSeq 6000 S4 Reagent Kit (Illumina, San Diego, California). Alignment was performed using DRAGEN (v.SW: 01.011.269.3.2.8) (Ma et al. 2015); data conversion from CRAM to BAM files with samtools (v.1.9); data pre-processing with Picard RevertSam, MarkDuplicates, SortSam, and MergeBamAlignment (v2.18.2) (Broad Institute 2019) and base quality recalibration with GATK BQSR (v.4.0.3.0). Germline variant calling and single-sample gVCF generation were performed using GATK HaplotypeCaller (v.3.5-0-g36282e4). Genotype analysis of trios/families, *de novo* variant discovery, variant quality recalibration, joint variant calling on all samples, and recalculation of genotype call accuracy on the population level were performed with GATK tools (Van der Auwera et al. 2013): HaplotypeCaller (v.3.5-0-g36282e4), VariantAnnotator (v.3.8-0-ge9d806836), VQSR, CalculateGenotypePosteriors, and VariantFiltration (v.4.0.12.0). All outputs were released to the DRC Portal and dbGaP, where the files can be accessed and downloaded (github.com/kids-first/kf-alignment-workflow).

High confidence *de novo* SNVs and indels were called for variants present only in the proband with GQ scores ≥ 20 for all trio members. Resulting variants were lifted to GRCh37/hg19 using UCSC’s liftOver tool and annotated using Annovar (v2013_09_11) (Wang et al. 2010). The liftover was necessary because the TraP and GWAVA annotations (see Variant prioritization) are not available under hg38.

### Variant prioritization

Variants were subset to include SNVs with gnomAD (Karczewski et al. 2020) minor allele frequency (MAF) < 1% (hereafter defined as rare). Next, we prioritized SNVs that:

1) were not present in gnomAD (novel);
2) affected a gene already known to cause a phenotype overlapping that of PHACE in OMIM (omim.org) or GWAS annotation (Buniello et al. 2019);
3) affected a gene where the mouse model phenotype (informatics.jax.org) overlapped that of PHACE;
4) affected genes disrupted by CNVs described by Siegel et al. (2013); or
5) affected genes with coding variants in two or more patients.

Prioritized coding variants were further evaluated for pathogenicity prediction scores using VarSome (Kopanos et al. 2019).

Prioritized noncoding variants were further evaluated for scores designed to predict effects on splicing [TraP (Gelfman et al. 2017), Human Splicing Finder (HSF) (Desmet et al. 2009)]; functional DNA [CADD (Rentzsch et al. 2019), DANN (Quang et al. 2015), GWAVA (Ritchie et al. 2014), EIGEN (Ionita-Laza et al. 2016), FATHMM (Shihab et al. 2015)]; transcription factor binding sites (TFBS) (Euskirchen et al. 2007); and open chromatin in vascular cells in Encode (Ernst et al. 2011).

Finally, we investigated the *de novo* indels identified in the genes prioritized using metrics 1-5.

### DECIPHER analyses

We first selected the pathogenic or likely pathogenic deletions and duplications described in DECIPHER (Firth et al. 2009) in individuals with hemangioma. We compared the genes affected by these CNVs with the genes prioritized in our cohort (described above) to identify other individuals in DECIPHER with hemangiomas and CNVs affecting our prioritized genes.

We then selected all pathogenic or likely pathogenic CNVs described in DECIPHER in individuals with facial hemangioma. The “facial hemangioma” feature was chosen to represent PHACE because it is the most common manifestation of this syndrome. We identified 47 DECIPHER individuals with facial hemangioma, with a total of 55 CNVs (17 gains, 38 losses) disrupting 5,703 genes; 1,191 of these genes were disrupted in at least two DECIPHER individuals. This subset of 1,191 genes was compared to the list of rare, *de novo* variants identified in our cohort of patients with PHACE to prioritize further candidates.

An enrichment analysis was performed for each gene identified by the second DECIPHER analysis. The DECIPHER pathogenic or likely pathogenic deletions disrupting the gene of interest in individuals with facial hemangioma were classified as “cases”. The DECIPHER pathogenic or likely pathogenic deletions disrupting the same gene in individuals without facial hemangioma were classified as “controls”. For each gene we built a contingency table comparing the number of cases to controls and performed a Fisher’s exact test (FET) using a cutoff of p-value < 0.05 for statistical significance. For significant genes we also calculated an odds ratio.

### Sanger sequencing

Prioritized candidate variants were validated by Sanger sequencing. DNA from the proband and each parent was amplified using Accuprime Taq polymerase (Invitrogen; 12339-016) using standard thermal cycling conditions with annealing at 60°C and variant-specific primer sequences (Supplemental Table 2). Small aliquots of each PCR product were run on a 1% agarose gel to verify amplification, and the remainder of each PCR product was cleaned up using a mixture of exonuclease 1 (Affymetrix; 70073X) and shrimp alkaline phosphatase (Affymetrix; 78390) and sequenced using the same primers.

### Protein structure analysis

The experimentally derived structure of THBS2 was obtained from RCSB PDB (Berman et al. 2000) structure 1YO8 (Carlson et al. 2005). The effect of the identified p.Asp859Asn variant in THBS2 was examined using DynaMut (Rodrigues et al. 2018), which predicts the effects of missense mutations on known PDB structures.

### Pathway analysis

All 4,320 genes with rare, *de novo* SNVs were analyzed with gProfiler (Raudvere et al. 2019) to determine if they had similar functions or affected the same pathways. Multiple testing-adjusted p-values (using the default g:SCS method in gProfiler) were reported, and 0.05 was used as the cutoff for statistical significance.

### Expression analysis

The chromatin accessibility atlas and gene expression data of the human developing heart was derived from nuclei isolated from human fetal hearts (Ameen & Sundaram et al., *unpublished*). Briefly, the Chromium 10X platform (Satpathy et al. 2019) was used to generate scATAC-seq data from three primary human fetal heart samples at 6, 8, and 19 weeks post-gestation. The scRNA-seq data from fetal hearts were analyzed from published studies (Asp et al. 2019; Miao et al. 2020; Suryawanshi et al. 2020) using the batch correction algorithm Harmony (Korsunsky et al. 2019), and then annotated using known cell-type-specific marker genes.

## Data Availability

Datasets related to this article are accessible through the Kids First Data Resource Portal (kidsfirstdrc.org) and/or dbGaP (ncbi.nlm.nih.gov/gap) accession phs001785.v1.p1.

## Abbreviations used

SNV: single nucleotide variant
CNV: copy number variant
ECM: extracellular matrix

## ACKNOWLEDGMENTS

We thank all the families for their participation in this study, and the PHACE Syndrome Community and Pediatric Dermatology Research Alliance (PeDRA) for support of this project. We acknowledge the support of the Children’s Research Institute at Children’s Wisconsin and the Genomic Sciences and Precision Medicine Center at Medical College of Wisconsin.

The results analyzed and shown here are based in part upon data generated by Gabriella Miller Kids First Pediatric Research Program (Kids First) projects phs001785.v1.p1. Kids First was supported by the Common Fund of the Office of the Director of the National Institutes of Health (commonfund.nih.gov/KidsFirst). The Hudson-Alpha Institute for Biotechnology was awarded a U24HD090744-01 to sequence structural birth defect cohort samples submitted by investigators through the Kids First program (1X01HL140519-01).

This study was supported by the NICHD, NHLBI, and NIAMS under award numbers 1X01HL14519, R03HD098526, and R01AR064258, respectively. The content is solely the responsibility of the authors and does not necessarily represent the official views of the National Institutes of Health. Additional support was received from a SID SUN grant, PHACE Foundation Canada, and the Children’s Research Institute.

This study makes use of data generated by the DECIPHER community. A full list of centers who contributed to the generation of the data is available from decipher.sanger.ac.uk and via email from. Funding for the project was provided by Wellcome. The individuals who carried out the original analysis and collection of the data bear no responsibility for the further analysis or interpretation of it in this paper.

## AUTHOR CONTRIBUTIONS (CREDIT)

Elizabeth S. Partan: Conceptualization, Data curation, Investigation, Visualization, Writing – original draft, Writing – review & editing

Francine Blei: Data curation, Writing – review & editing

Sarah L. Chamlin: Data curation, Writing – review & editing

Olivia M. T. Davies: Data curation, Writing – review & editing

Beth A. Drolet: Data curation, Writing – review & editing

Ilona J. Frieden: Data curation, Writing – review & editing

Ioannis Karakikes: Writing – review & editing

Chien-Wei Lin: Formal analysis, Writing – review & editing

Anthony J. Mancini: Data curation, Writing – review & editing

Denise Metry: Data curation, Writing – review & editing

Anthony Oro: Writing – review & editing

Nicole S. Stefanko: Data curation, Writing – review & editing

Laksshman Sundaram: Investigation, Visualization

Monika Tutaj: Data curation, Formal analysis, Writing – review & editing

Alexander E. Urban: Writing – review & editing

Kevin C. Wang: Writing – review & editing

Xiaowei Zhu: Writing – review & editing

Nara Sobreira: Conceptualization, Funding acquisition, Project administration, Supervision, Writing – review & editing

Dawn H. Siegel: Conceptualization, Data curation, Funding acquisition, Project administration, Supervision, Writing – review & editing

