## Supplemental Tables for "Analysis of Whole Genome Sequencing in a Cohort of Individuals with PHACE Syndrome Suggests Dysregulation of RAS/PI3K Signaling"

**Supplemental Table 1: Full list of candidate *de novo* germline variants**

| Gene | Patient ID | Patient phenotype (HPO terms) | Variant (hg38) | Variant consequence | gnomAD MAF | Predicted effect on splicing? | Damaging functional DNA scores? | TFBS? | Open chromatin in HUVECs? | OMIM phenotype | Homozygous knockout mouse model | CNV(s) |
| --- | --- | --- | --- | --- | --- | --- | --- | --- | --- | --- | --- | --- |
| *AFF2* | Patient 7 | HP:0001028, HP:0000329, HP:0100545, HP:0005116, HP:0005344, HP:0100659, HP:0001629, HP:0001655, HP:0011590, HP:0012020, HP:0010775 | X:148547549:  A:G | intronic SNV (intron 1) | novel | — | — | — | — | mental retardation, FRAXE type | — | 1 duplication in PHACE |
|  | Patient 2 | HP:0001028, HP:0000329, HP:0005116, HP:0002616, HP:0002631 | X:148758931:  G:A | intronic SNV (intron 3) | novel | — | — | — | — |  |  |  |
| *ALDH3A1* | Patient 8 | HP:0001028, HP:0100545, HP:0005116, HP:0005344, HP:0100659 | 17:19739095:  C:T | p.Val373Met | novel | HSF: changes exonic splicing silencer / enhancer | CADD=23.3; FATHMM=0.48 | POLR2A, MYC | — | — | no phenotype^[1]^ | 1 deletion and 1 duplication in DECIPHER [IDs: 399186^[2]^, 399297] |
| *ATP7B* | Patient 10 | HP:0001028, HP:0000329, HP:0100545, HP:0005116, HP:0005344, HP:0100659, HP:0001655 | 13:51970591:  T:C | p.Thr482Ala | 1.2e-5 | HSF: activates cryptic acceptor site | — | — | — | Wilson disease | copper accumulation; liver cirrhosis^[3]^ | 1 deletion and 1 duplication in DECIPHER [IDs: 395925, 253898] |
| *BCAS3* | Patient 15 | HP:0001028, HP:0000329, HP:0100545, HP:0100659, HP:0001655 | 17:61029145:  A:T | intronic SNV (intron 17) | novel | TraP=0.457; HSF: activates cryptic donor site | EIGEN=0.26 | — | — | — | abnormal cardiovascular patterning; embryonic lethality^[4]^ | 1 deletion in DECIPHER [ID: 395912] |
|  | Patient 18 | HP:0001028, HP:0000329, HP:0012020, HP:0000365, HP:0000405, HP:0000682 | 17:61087142:  G:A | intronic SNV (intron 23) | 3.2e-5 | HSF: activates cryptic acceptor site, changes exonic splicing silencer / enhancer | CADD=12.54; FATHMM=0.84; EIGEN=0.53 | GATA1 | — |  |  |  |
|  | Patient 20 | HP:0001028, HP:0000329, HP:0100545, HP:0005344 | 17:61370924:  CTT:C | intronic 2-bp deletion (intron 24) | 7.0e-6 | — | — | — | — |  |  |  |
| *DLC1* | Patient 1 | HP:0001305, HP:0001317, HP:0007033, HP:0001320, HP:0001321, HP:0001028, HP:0000329, HP:0100545, HP:0005116, HP:0005344, HP:0100659 | 8:13089422:  C:T | intronic SNV (intron 15) | 1.9e-3 | — | — | — | transcriptional elongation | colorectal cancer, somatic | neural tube, heart, brain, and vascular placenta defects; embryonic lethality^[5]^ | 1 duplication in DECIPHER [ID: 395935] |
|  | Patient 17 | HP:0001035, HP:0001028, HP:0000329, HP:0002564, HP:0001680 | 8:13104433:  G:A | intronic SNV (intron 7) | novel | — | EIGEN=0.25 | — | transcriptional elongation |  |  |  |
|  | Patient 3 | HP:0001028, HP:0000329, HP:0100545, HP:0005116, HP:0005344, HP:0100659 | 8:13253055:  G:C | intronic SNV (intron 5) | novel | HSF: activates cryptic acceptor site | — | — | strong enhancer |  |  |  |
| *EPHA3* | Patient 9 | HP:0001028, HP:0000329, HP:0001643, HP:0000682 | 3:89246570:  G:A | intronic SNV (intron 3) | novel | HSF: activates cryptic acceptor site, changes exonic splicing silencer / enhancer | EIGEN=0.58 | CTCF, TCF12, RAD21 | — | — | perinatal death due to cardiac failure^[6]^ | 1 duplication in PHACE |
| *EXOC4* | Patient 6 | HP:0001317, HP:0001320, HP:0001028, HP:0000329, HP:0100545, HP:0005116, HP:0005344, HP:0100659, HP:0001655, HP:0004942, HP:0011611, HP:0001680 | 7:133264954:  G:A | intronic SNV (intron 1) | novel | HSF: activates cryptic acceptor site, changes exonic splicing silencer / enhancer | — | — | — | — | embryonic abnormalities; incomplete gastrulation; abnormal mesoderm formation; embryonic lethality^[7]^ | 1 deletion in PHACE |
|  | Patient 19 | HP:0001028, HP:0000329 | 7:133538908:  G:A | intronic SNV (intron 9) | novel | HSF: changes exonic splicing silencer / enhancer | — | — | — |  |  |  |
|  | Patient 17 | HP:0001035, HP:0001028, HP:0000329, HP:0002564, HP:0001680 | 7:133931491:  A:G | intronic SNV (intron 13) | novel | HSF: activates cryptic acceptor and donor sites | — | — | — |  |  |  |
|  | Patient 13 | HP:0001305, HP:0001028, HP:0000329, HP:0100545, HP:0005344, HP:0001629, HP:0001655, HP:0000365, HP:0000407, HP:0000682 | 7:134017984:  G:A | intronic SNV (intron 17) | 9.6e-5 | HSF: activates cryptic donor site | EIGEN=0.05; GWAVA=0.5 | GATA2, GATA3 | — |  |  |  |
| *GLRX3* | Patient 11 | HP:0001028, HP:0000329, HP:0100545, HP:0005116, HP:0005344, HP:0100659 | 10:130174209:  C:T | intronic SNV (intron 8) | novel | TraP=0.498; HSF: activates cryptic donor site | — | — | transcriptional elongation | — | pericardial effusions; embryonic lethality^[8]^ | 1 deletion in DECIPHER [ID: 396151] |
| *PIK3CA* | Patient 5 | HP:0001028, HP:0000329, HP:0001655, HP:0001643 | 3:179200365:  T:G | intronic SNV (intron 3) | novel | HSF: changes exonic splicing silencer / enhancer | EIGEN=0.21 | — | weakly transcribed | CLAPO syndrome, somatic; CLOVE syndrome, somatic; Cowden syndrome; keratosis, seborrheic, somatic; macrodactyly, somatic; megalencephaly-capillary malformation-polymicrogyria syndrome, somatic; nevus, epidermal, somatic; variety of cancers, somatic | vascular defects; hemorrhage; growth retardation; embryonic lethality^[9]^ | 1 duplication in PHACE; 1 duplication in DECIPHER [ID: 283584] |
| *RASA3* | Patient 4 | HP:0001317, HP:0001320, HP:0001028, HP:0000329, HP:0100545, HP:0005116, HP:0005344, HP:0100659, HP:0000609, HP:0000682 | 13:114041023:  C:T | p.Val85Met | 1.2e-5 | HSF: changes exonic splicing silencer / enhancer | CADD=23.6; FATHMM=0.95; GWAVA=0.44 | — | transcriptional elongation | — | hemorrhage; embryonic lethality^[10]^ | 1 deletion in DECIPHER [ID: 393047] |
| *STAMBP* | Patient 16 | HP:0001028, HP:0000329, HP:0100545, HP:0005344, HP:0000365, HP:0000405, HP:0000682 | 2:73850410:  T:A | p.Ile301Asn | novel | — | CADD=27.7; FATHMM=0.99 | — | weakly transcribed | microcephaly-capillary malformation syndrome | postnatal growth retardation; limb-clasping; hypocellular cerebral cortex; loss of hippocampal neurons; blepharoptosis; death of starvation at weaning^[11]^ | — |
| *THBS2* | Patient 14 | HP:0001317, HP:0007033, HP:0001321, HP:0001028, HP:0000329, HP:0100545, HP:0005116, HP:0005344, HP:0100659 | 6:169225343:  C:T | p.Asp589Asn | novel | — | CADD=28.4; FATHMM=0.96 | — | — | — | blood vessel abnormalities; premature death^[12]^ | 5 deletions in DECIPHER [IDs: 392300^[13]^, 392970^[14]^, 392995^[15]^, 396392^[16]^, 402744] |
|  | Patient 12 | HP:0001028, HP:0000329, HP:0100545, HP:0005116, HP:0005344, HP:0100659, HP:0001629, HP:0001655 | 6:169243142:C: CCACATTCT | intronic 8-bp insertion (intron 5) | novel | — | — | IKZF1 | — |  |  |  |
|  |  |  | 6:169243176:  C:T | intronic SNV (intron 5) | 3.3e-5 | — | FATHMM=0.41 | IKZF1 | — |  |  |  |

**Supplemental Table 2: Primers for Sanger sequencing**

| Patient ID | Locus | Forward primer sequence | Reverse primer sequence | Amplicon size (bp) |
| --- | --- | --- | --- | --- |
| Patient 1 | *DLC1* intron 15 | CAGCCTGGGTGATGAGAATG | TGGTCAGAATCCCTTTGCAC | 688 |
| Patient 2 | *AFF2* intron 3 | CCCTTTGGCAGACATTTCCT | ACATATTCTTGGCTGGGTGC | 710 |
| Patient 3 | *DLC1* intron 5 | TGACACAGGAAAGGGAGGAA | GAACGAGGACATCCAATGACA | 541 |
| Patient 4 | *RASA3*–p.Val85 | AGGCAAAGGGAAAACCCAAA | CGTGGTGAAGTCTTCTCTGC | 979 |
| Patient 5 | *PIK3CA* intron 3 | TGCAGTCTTCTACCTGTGTCT | AGGTTTGGTTGTTCACAGTACA | 785 |
| Patient 6 | *EXOC4* intron 1 | ACAGGCTTTTGTTTCCCTGAT | TGGGCCCCTAAGTGGATAAA | 650 |
| Patient 7 | *AFF2* intron 1 | GATCCCTTTGTCTGGTTGCA | TTGACTTGCAGTGGGAAGAC | 673 |
| Patient 8 | *ALDH3A1*–p.Val373 | GGTGGTCCCTCCTGAATTTG | ATCCAGTTCATCAACCAGCG | 657 |
| Patient 9 | *EPHA3* intron 3 | TCTTTGACCCTCCCACTAGG | TTCCCAGTCTCAGAGGATGG | 710 |
| Patient 10 | *ATP7B*–p.Thr482 | TACGAGGTCTATACGCAGCA | GCCAGCGTGATTTGTTTCTC | 682 |
| Patient 11 | *GLRX3* intron 8 | TTGGGAATTGCACCTGTACC | CTCAGTTCTGCTCTCTGCTG | 584 |
| Patient 12 | *THBS2* intron 5 (2 variants) | GGCTCCTTGCTCTGCTTTAA | ACGATCTAGGGCTTCAGAGG | 799 |
| Patient 13 | *EXOC4* intron 17 | ATTTTCCTAGGGGCCAGAGT | CCCTTCCCTGGCAGAAAAAT | 800 |
| Patient 14 | *THBS2*–p.Asp859 | AGAAGCCATTTTCCTCCACG | TTTCCAGCGAAGAAGCACAT | 549 |
| Patient 15 | *BCAS3* intron 17 | GTTACTTTCCCAGTGGTGCA | CCAGGGAAGTCCAAAGTGAC | 533 |
| Patient 16 | *STAMBP*–p.Ile301 | TACCTTTCCACTGTCGGGAT | ACATTACCAGAAGGCTGCAC | 726 |
| Patient 17 | *DLC1* intron 7 | GAGTCCTTGCACAGAATGGAA | CAGGAAAATTGGGGCCTTGA | 428 |
|  | *EXOC4* intron 13 | GGTGCCTAGCCGGTTTTATT | CACATCACCCTTATGGAGGC | 583 |
| Patient 18 | *BCAS3* intron 23 | GATGGCATTGACCCTCTCTG | AAGCAACTTCCTGATGTGCA | 594 |
| Patient 19 | *EXOC4* intron 9 | TTGACCCCAGGAGTTAGAGG | AGGTACAAGACACCTGGGTAA | 339 |
| Patient 20 | *BCAS3* intron 24 | CTTGCTCTGTCACCAGACTG | ATTGCTGTGTCACATGTGGT | 684 |
